# Caesarean myomectomy is not a risk factor for blood products transfusion among pregnant individuals with fibroids: a retrospective cohort study

**DOI:** 10.1101/2024.09.24.24314170

**Authors:** S Jarvi, A Aviram, C Joliffe, S Wortsman, GY Liu, A Berndl, J Barrett, J Kroft

## Abstract

**Background:** Uterine fibroids are common benign tumors during reproductive age. Among pregnant individuals with uterine fibroids, myomectomy at the time of cesarean delivery (CD) allows individuals to avoid a second surgery later in life.

**Objective:** To examine the safety of myomectomy performed at the time of CD and the factors that increase the risk of greater blood loss in cesarean myomectomy.

**Study design:** This was a retrospective cohort study of all cesarean myomectomies (CM) performed at a tertiary care center in Toronto, Ontario between January 1, 2012, and June 1, 2019. The study group included individuals who had CM, and the control group included individuals with fibroids who had a CD without removing their fibroids. The primary outcome was the rate of blood products transfusion. Secondary outcomes were the need for hysterectomy and estimated blood loss (EBL). Subgroup analyses were performed for fibroid size, previous CD, and elective vs urgent CD. Logistic regression analysis was performed to assess for confounders.

**Results:** Overall, 110 CM were compared with 152 CD. The overall transfusion rate for CM was 7.3% compared to 0.7% in the control group (p = 0.005). All procedures requiring transfusion from both groups were performed < 37^0/7^ weeks of gestation. In logistic regression analysis, performing a myomectomy was not associated with a higher risk for transfusion. The only factors associated with transfusion were preterm birth (aOR 14.43, p=0.049) and low pre-operative hemoglobin (aOR 0.90, p=0.02). The median reported EBL was 800 mL for CM and 700 mL for controls (p<0.001). No individuals in either group required concomitant hysterectomy. Subgroup analysis showed that fibroid size ≥5cm and previous CD were not associated with increased risk of transfusion among those who had a CM, while urgent CM was significantly associated with the need for transfusion (p=0.02).

**Conclusion:** This study demonstrates that the transfusion rate was not significantly higher in those undergoing CM at term, compared to those with fibroids undergoing CD without a myomectomy.

## INTRODUCTION

Uterine fibroids are common benign tumors, that originate from the smooth muscle layer of the uterus^1^. It is estimated that fibroids occur in over 70% of biologically female individuals by the onset of menopause^2^. Individuals with fibroids are often asymptomatic, however, symptoms may include uterine bleeding, pelvic pain, pressure, and infertility^3^. There are multiple treatment options including medical, surgical, or image-guided. Myomectomy and hysterectomy are the mainstays of surgical treatment. While hysterectomy is a definitive treatment, myomectomy is reserved for those individuals wishing to preserve fertility.

Fibroids are also common among pregnant individuals and are becoming more frequent as maternal age increases. While they are often asymptomatic, they may cause pain, pressure, or preterm labor. If left untreated, many of these individuals will require surgical treatment later in life, and it is estimated that treatment will be required in 25% of individuals with fibroids^2^. Cesarean delivery (CD) presents a unique opportunity to simultaneously perform a myomectomy with the CD, possibly preventing a second surgery in the future. Cesarean myomectomy (CM), however, is controversial. It has historically been discouraged because of the risk of excessive hemorrhage causing anemia or requiring blood products transfusion, the risk of unnecessary hysterectomy, puerperal infection, and increased operative time^4^. However, more recent literature has supported CM to be a safe procedure with only a small increase in operative time and no increased risk of post-operative complications^5,6^. Furthermore, there may be benefits of doing a CM, such as a smaller uterine incision because the uterus/fibroid ratio is increased and the fibroid cleavage plans are easier to find^7^. As well, the natural uterine contractibility and involution of the postpartum period reduces hemorrhage^8^.

Although recent systematic reviews and meta-analyses of observational studies have provided favorable evidence to support carrying out CM, the data is heterogeneous in terms of the control groups and types of fibroids removed and there is potential bias from the included studies.

The purpose of this study was to therefore compare the surgical outcomes of individuals undergoing CM compared to those with fibroids undergoing CD alone.

## METHODS

We conducted a single-center retrospective cohort study of all CM performed between January 1, 2012, and June 1, 2019. A control group of individuals that underwent CD in which fibroids were identified but not removed was evaluated over a similar period. These individuals were identified from the hospital database and data was extracted from patient records.

Extracted data included demographics such as maternal age (years), parity, gestational age at delivery (determined by first trimester ultrasound), previous myomectomy, and size and number of fibroids removed. The primary outcome was the rate of blood products transfusion. Secondary outcomes were the need for hysterectomy, length of surgery (minutes) and estimated blood loss (EBL, mL, determined subjectively by the surgical team), and change in hemoglobin (Hb) from pre-operative (within one month of surgery) to post-operative (day 1 after surgery). Individuals with a history of a documented bleeding disorder, multiple gestation, pre-eclampsia, chorioamnionitis, placenta previa, placenta accrete spectrum disorders, or abruption were excluded. Subgroup analyses were performed comparing outcomes for individuals depending on fibroid size (≥ or < 5cm), previous CD, and elective vs urgent CD.

Statistical analysis was performed using SPSS version 28.0 (IBM, Armonk, NY, USA). Continuous variables were analyzed using the Mann-Whitney U test, and categorical variables using the chi-square or Fisher’s exact test, as needed. A logistics regression analysis was performed to account for potential confounders, and a sub-group analysis was performed as well among the individuals who had a myomectomy. The significance level was determined at p < 0.05. This study was approved by the hospital Research Ethics Board (Project Identification Number: 212-2019).

## RESULTS

Overall, 262 individuals met the inclusion criteria and were included in the study: 110 (42%) who underwent a myomectomy during their cesarean delivery (CM group), and 152 (58%) who had fibroids that were not removed during their CD (CD group).

The participants in the CM group were characterized by a higher rate of previous surgery for myomectomy, slightly lower gestational age at the time of delivery, and a higher rate of elective procedure. No other demographic characteristics were found to be different between the groups (Table 1).

**Table 1.**
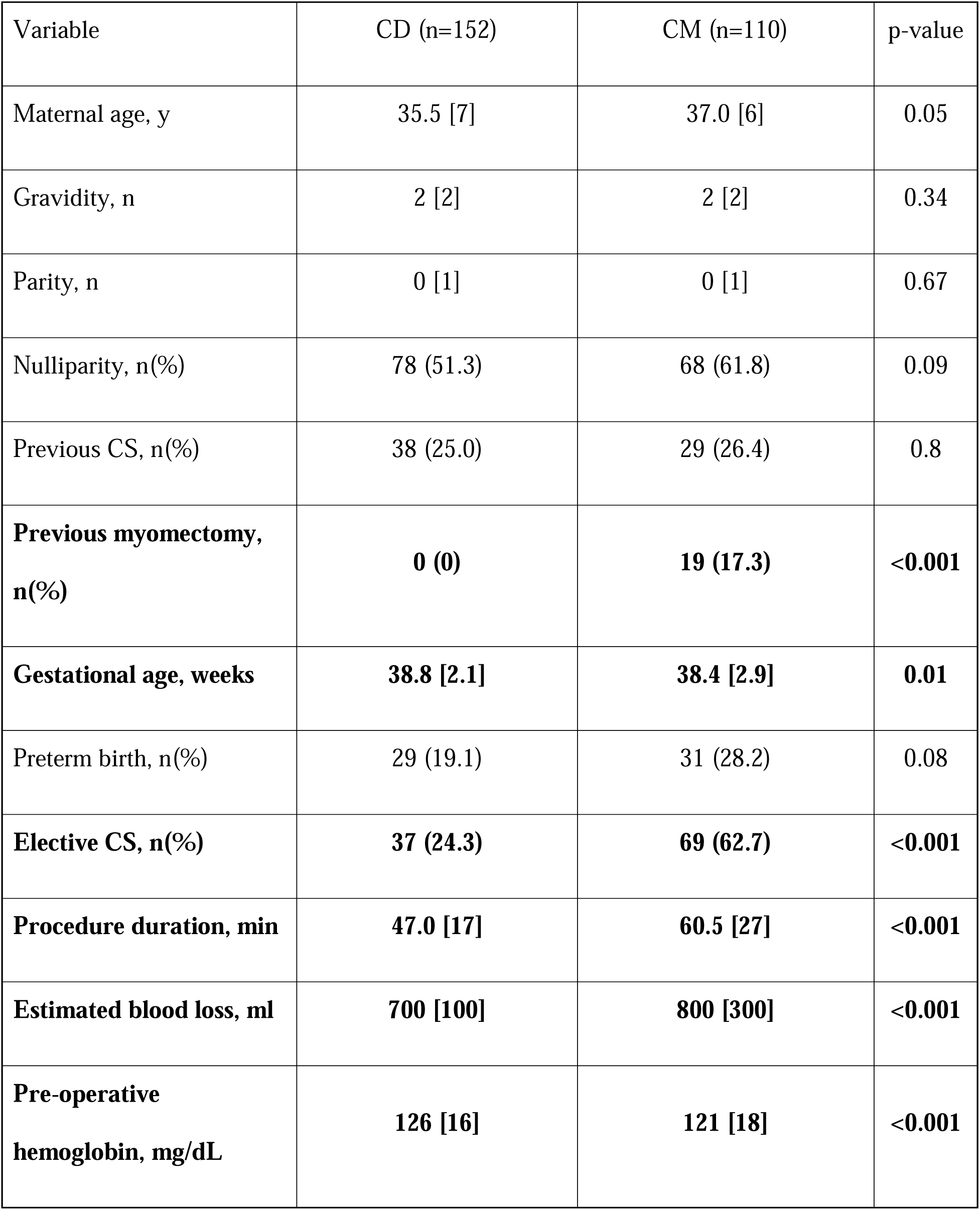
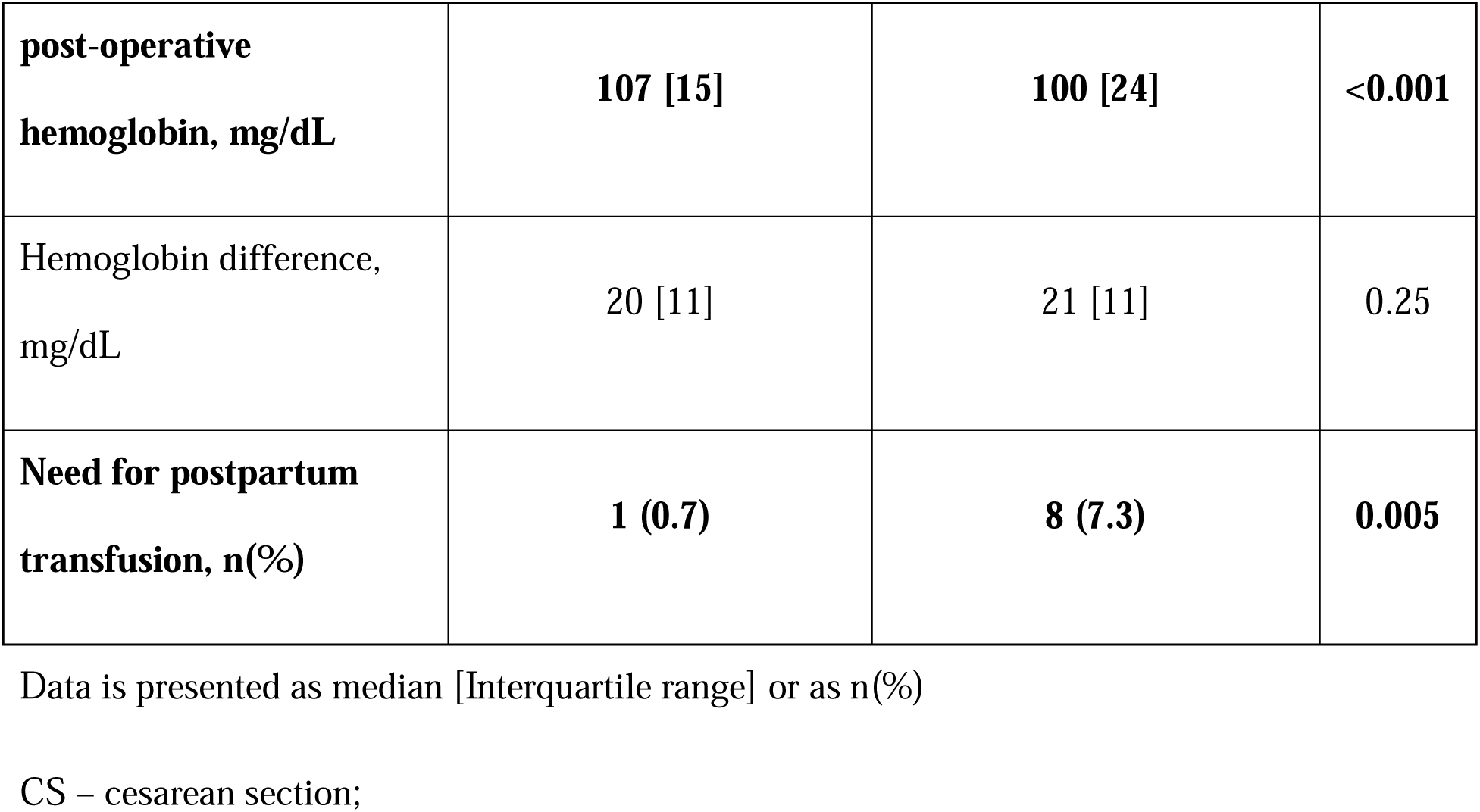
Characteristics.

As for the procedures, CM took approximately 13.5 minutes longer to complete compared with regular CD (47.0 vs 60.5 minutes, p<0.001). The estimated blood loss was higher in the CM group, as was the rate of post-procedure need for blood products transfusion (Table 1). Yet, the difference between the hemoglobin level before and after the procedure was not significantly different between the groups (21 mg/dL in the CM group vs. 20 mg/dL in the CD group, p=0.25).

To account for potential confounders for the need for post-procedure transfusion, a logistic regression model was calculated using the following factors: Myomectomy, pre-operative hemoglobin level, preterm birth, maternal age, previous surgery for myomectomy, previous CD and urgent vs. non-urgent procedure (Table 2). Pre-operative hemoglobin level and preterm birth were found the be independent risk factors for transfusion, but myomectomy during the CD was not (aOR 5.4. 95% CI 0.47-61.6, p=0.18).

**Table 2.** logistic regression, risk factors for transfusion.

| Variable | Adjusted odds<br>ratio | 95% confidence<br>interval | p-value |
| --- | --- | --- | --- |
| Myomectomy | 5.39 | 0.47-61.6 | 0.18 |
| **Pre-operative hemoglobin (for every**<br>**increase in 1mg/dL)** | **0.90** | **0.82-0.99** | **0.02** |
| **Preterm birth** | **14.43** | **1.01-206.93** | **0.049** |
| Maternal age (every additional year) | 0.95 | 0.75-1.18 | 0.62 |
| Previous myomectomy | 5.77 | 0.37-90.99 | 0.21 |
| Previous CS (each additional CS) | 1.21 | 0.23-6.52 | 0.82 |
| Non-elective CS | 3.13 | 0.17-57.55 | 0.44 |

### Size of fibroid

Out of the 110 individuals who had a myomectomy, 53 (48%) had a fibroid size less than 5 cm (<5cm), and 57 (52%) had a fibroid size greater or equal to 5 cm (≥5cm). Those with larger fibroid size were characterized by a lower rate of previous surgery for myomectomy, their CM took approximately 8 minutes longer, and their estimated blood loss was higher, compared with the smaller fibroid group. However, the hemoglobin difference before and after the procedure, and the rate of postpartum transfusion were not significantly different between the groups (Table 3a).

**Table 3a.** characteristics, only myomectomies, comparison based on size.

| Variable | Fibroid < 5cm (n=53) | Fibroid ≥ 5cm (n=57) | p-value |
| --- | --- | --- | --- |
| **Procedure duration, min** | **57 [21]** | **65 [27]** | **0.03** |
| **Estimated blood loss, ml** | **700 [100]** | **900 [300]** | **0.001** |
| Pre-operative hemoglobin, mg/dL | 121 [21] | 121 [15] | 0.68 |
| post-operative hemoglobin, mg/dL | 100 [25] | 100 [21] | 0.36 |
| Hemoglobin difference, mg/dL | 18 [14] | 21 [16] | 0.16 |
| Need for postpartum transfusion, n(%) | 3 (5.7) | 5 (8.8) | 0.53 |

Data is presented as median [Interquartile range] or as n(%)
No difference in other characteristics, except for previous myomectomy (26.4% in the small
fibroid group, and 8.8% in the large fibroid group)

### Previous cesarean section (CS)

Overall, of the individuals with CM, 81 individuals (74%) had no previous CD and 29 (26%) had at least one previous CD. There was no significant difference in the hemoglobin decline from before to after the procedure, or in the rate of postpartum transfusion between the groups (Table 3b).

**Table 3b.** characteristics, only myomectomies, comparison based on previous CS.

| Variable | No previous CS (n=81) | Previous CS (n=29) | p-value |
| --- | --- | --- | --- |
| Procedure duration, min | 60 [27] | 61 [27] | 0.83 |
| **Estimated blood loss, ml** | **800 [300]** | **700 [275]** | **0.02** |
| Pre-operative hemoglobin, mg/dL | 121 [16] | 125 [22] | 0.26 |
| **post-operative hemoglobin, mg/dL** | **97 [21]** | **105 [21]** | **0.03** |
| Hemoglobin difference, mg/dL | 21 [14] | 20 [18] | 0.10 |
| Need for postpartum transfusion, n(%) | 3 (5.7) | 5 (8.8) | 0.53 |

Data is presented as median [Interquartile range] or as n(%)
No difference in other characteristics, except for preterm birth rate (34.6% in the no previous CS group vs. 10.3% in the previous CS group) and elective procedure (50.6% in the no previous CS group vs. 96.6 in the previous CS group).

### Urgent versus elective procedure

Of the individuals with CM, 41 (37%) participants had an urgent procedure and 69 (63%) had an elective procedure. The urgent CM group was characterized by a higher rate of primiparity and a lower gestational age at delivery (Table 3c). The procedure duration was not significantly different between the two groups. The pre-operative hemoglobin and hemoglobin differences from before to after surgery were also not significantly different between the two groups. However, the estimated blood loss was higher by approximately 100mL in individuals with urgent CM, the post-operative hemoglobin was lower by approximately 10mg/dL, and the rate of post-procedure blood transfusion was higher among individuals with urgent CM (14.6% in urgent CM group vs 2.9% in elective CM group, p=0.02) (Table 3c).

**Table 3c.** characteristics, only myomectomies, comparison based on elective vs. urgent.

| Variable | Urgent (n=41) | Elective (n=69) | p-value |
| --- | --- | --- | --- |
| Procedure duration, min | 58 [31] | 62 [24] | 0.43 |
| **Estimated blood loss, ml** | **900 [400]** | **800 [225]** | **0.02** |
| Pre-operative hemoglobin, mg/dL | 118 [17] | 122 [17] | 0.16 |
| **post-operative hemoglobin, mg/dL** | **94 [18]** | **104 [23]** | **0.02** |
| Hemoglobin difference, mg/dL | 23 [19] | 20 [17] | 0.15 |
| **Need for postpartum transfusion, n(%)** | **6 (14.6)** | **2 (2.9)** | **0.02** |

Data is presented as median [Interquartile range] or as n(%)
No difference in other characteristics, except for parity (78% of primiparous individuals in the urgent group vs. 52.2% in the elective group) and gestational age (34.9 weeks in the urgent group vs. 38.9% in the elective group)

## DISCUSSION

In this cohort of pregnant individuals with fibroids undergoing cesarean delivery, 42% had a concurrent myomectomy at the time of delivery. Participants who underwent CM had a slightly longer procedure duration than their counterparts who did not have a myomectomy during the CD. Individuals in the CM group had a higher rate of post-procedure need for blood products transfusion. However, when accounting for other factors that may impact the need for post-procedure transfusion, the performance of a myomectomy was not found to be a risk factor for transfusion. Fibroid size (fibroid size <5 cm versus ≥ 5cm) and previous CD were not found to impact post-procedure transfusion rates, while urgent (vs. elective) procedure was associated with a higher rate of transfusion.

In our study, the CM group had a longer procedure duration of 13.5 minutes compared to the CD. In systematic reviews and primary studies, procedure time was found to be longer in CM than in CD alone^5,6^. In a meta-analysis of 23 studies, the procedure duration was longer in CM by 10.4 minutes, and in another meta-analysis (17 studies) it was on average 14.8 minutes longer which is consistent with our study^5,6^. This increased surgical time did not translate to a clinically meaningful complication, and compared with the potential alternative, which is another surgical procedure, is considerably more cost-effective in terms of operative room time, personnel, etc.

We found the mean decline in hemoglobin was not significantly different among participants who had CM vs. CD. This is consistent with several retrospective cohort studies in the literature that have also found no significant difference in the hemoglobin decline between CM and CD in individuals with fibroids^4,9–16^. However, two recent meta-analyses found that the mean change in hemoglobin was of greater magnitude among CM compared to CD by a clinically insignificant amount (0.27 gm% and 0.20g/dL differences between CM and CD individuals)^5,6^. The larger sample sizes in these studies (n=3960 and 4906 individuals respectively) could explain why a statistically significant difference was found with a small effect size.

A large retrospective cohort study examining predictors of peripartum blood transfusion found that earlier gestational age and pre-operative anemia (hematocrit <32%) were both significantly associated with an increased odds of needing transfusion at the time of CD alone^17^. Several retrospective cohort studies have found no difference in transfusion rates between CM and CD ^1,4,10,12,15,16,18,19^. Two studies reported that myomectomy was not associated with an increased risk of blood loss, greater decline in hemoglobin, or increased risk of postpartum hemorrhage during CD^1,9^. In contrast, two recent meta-analyses found a higher rate of transfusion among CM compared to CD (OR 1.47 and RR 1.45 respectively)^5,6^. However, the outcomes of these meta-analyses may be confounded by the factors identified in our study of pre-operative hemoglobin and preterm birth.. So, although CM may appear to be associated with a higher rate of transfusion in some studies, this association could be due to other factors independent of myomectomy, which was highlighted in our study, such as pre-operative anemia and preterm birth.

Fibroid size has been studied as a factor impacting the outcomes of CM. In our study, among CM individuals, a larger fibroid size (≥5 cm) was associated with longer procedure duration and higher estimated blood loss by 200mL, but no significant difference in transfusion rate. Several studies have found that larger fibroid diameter was associated with increased blood loss events such as postpartum hemorrhage^1,9,18^, while other studies have found no significant difference in transfusion rate or hemoglobin decline^4,14–16^. The heterogeneity in results may be due to different cutoff points for fibroid size in different studies, as well as the experience of the surgeons with performing CM on large fibroids. In one retrospective cohort study by Lee, no significant difference was found in transfusion rate comparing myomectomy for fibroids <5cm and 5-10 cm, but a significantly higher rate of transfusion in individuals with fibroids >10cm was found^20^. Also, many studies did not specify whether the control group undergoing CD had fibroids, which would lead to a bias in the results. In one retrospective study, the presence of fibroids larger than 7cm increased the rate of massive intra-operative bleeding regardless of whether myomectomy was performed.^21^

The higher transfusion rate among urgent (versus elective) CM individuals in our analysis could be explained by other confounding factors, since it was not found to be a predictor of transfusion in the logistic regression comparing CD with CM. A retrospective cohort study found that unplanned CD in the first stage of labor was not significantly associated with EBL>500mL or a hemoglobin decline >20g/L in multivariate logistic regression analysis of individuals with fibroids who underwent CD or CM^9^. However, it is possible that a non-elective procedure could be a predictor in the CM group. So, although we cannot make any conclusions from this sub-group analysis, it is possible that an urgent CM could be a higher risk for transfusion. This should be documented in future studies as a potential confounding variable and surgeons should be prepared for this possibility when undertaking urgent cesarean section that may necessitate a myomectomy concurrently.

This study identified preterm birth and pre-operative hemoglobin as significant predictors of the risk of needing transfusion among CD and CM individuals with fibroids. Pre-operative hemoglobin levels should be optimized by clinicians throughout pregnancy to decrease the risk of peripartum transfusion. Preterm birth cannot necessarily be modified and therefore in cases of preterm birth and known myomectomy, physicians should have blood products available as the risk of transfusion is higher. Additional precautions for hemostasis can be taken with individuals who have fibroids and may require a CM, similar to methods used during abdominal myomectomy such as uterine tourniquet^22^. We also found that when accounting for these factors, myomectomy itself was not significantly associated with an increased risk of transfusion. This contributes to the growing body of literature that is turning in favor of CM as a safe procedure that does not cause additional blood loss risks.

There is heterogeneity in the literature on whether fibroid size impacts transfusion rate; ultimately this may be a factor that varies based on the center and surgical expertise. Also, fibroid location and blood conservation techniques can greatly impact blood loss and the need for transfusion, as seen in the gynecology literature^5,22^. Therefore, it is not surprising that there is heterogeneity in previous studies. This remains an unanswered question and future systematic reviews should address this with a subgroup analysis on fibroid sizes <5 cm compared to ≥5 cm as these appear to be the categories used by most of the studies we found.

Future work should also concentrate on the adaptation of methods used to decrease intraoperative blood loss that are currently used for myomectomy outside of pregnancy. Several strategies have been identified to decrease intraoperative blood loss at the time of myomectomy: temporary uterine artery occlusion with suture ligation, vascular clips, or peri-cervical tourniquet; uterotonics, fibroid dissection with electrosurgical devices; and use of antifibrinolytic agents.^14,16,23,24^ Use of a cell saver may also be considered if anticipating a large amount of blood loss. It will be important to study these methods at the time of CS to determine if they are adaptable from the gynecology literature.

One of the strengths of our study is that both groups had fibroids, in contrast to most previous studies in which a comparison was made between individuals with fibroids who had a myomectomy and those without fibroids who had a CD. Additionally, we included individuals with large fibroids >5 cm; the largest fibroid included was 30cm in diameter. We also did a subgroup analysis on the impact of previous CD and urgent CD on the outcomes of CM, which we have only seen in one other study^9^. Finally, not all studies included intramural fibroids, while our study included all types of fibroid locations. We also had 21 different surgeons in our center carrying out CM which makes the results more generalizable compared to other studies where only the most experienced surgeons may have been carrying out CM or it was unknown.

The main limitation of our study is its retrospective nature. We also lacked data on fibroid size among the control individuals. Another limitation is the lack of data on long-term outcomes, such as hospitalization duration, fibroid recurrence, uterine dehiscence, and post-surgical adhesions. Also, fibroid location (intramural, subserosal, pedunculated) was not accounted for in our study and was not described in many of the previous studies, which can bias the results. An important aspect of performing CM is the decision-making surrounding proceeding with an elective opportunistic myomectomy. The size of the fibroid, location, and blood loss during the procedure may impact whether the decision is made to go ahead with the myomectomy after the delivery of the infant. Therefore, caution must be taken not to generalize the results of our study and others to all CMs, as it is prudent to take all of the fibroid and patient characteristics into account before proceeding with an opportunistic CM.

## CONCLUSION

In our study, there was no significant difference in transfusion rate between CD and CM once other factors were accounted for. Preterm birth and pre-operative hemoglobin were the only significant predictors of needing a transfusion. Larger fibroid size and previous CD did not significantly impact transfusion rate among CM individuals, while urgent CD did. Overall, CM is a safe procedure compared to CD in terms of blood loss, but pre-operative hemoglobin should be optimized and blood conservation techniques utilized when possible, to decrease risk. CM offers many advantages including avoiding a second surgery, a potentially smaller incision on the uterus, better fibroid cleavage planes, and increased uterine contractility. Surgeon judgment and appropriate skills need to be utilized to minimize any increased risks to the patient.

## Data Availability

All data produced in the present study are available upon reasonable request to the authors

## Acknowledgements

None

## REFERENCES

1. Zhao R, Wang X, Zou L, Zhang W. Outcomes of Myomectomy at the Time of Cesarean Section among Pregnant Women with Uterine Fibroids: A Retrospective Cohort Study. Biomed Res Int. 2019;2019:1–6. doi:10.1155/2019/7576934

2. Stewart E, Cookson C, Gandolfo R, Schulze-Rath R. Epidemiology of uterine fibroids: a systematic review. BJOG An Int J Obstet Gynaecol. 2017;124(10):1501–1512. doi:10.1111/1471-0528.14640

3. Bedaiwy MA, Janiszewski P, Singh SS. A Patient Registry for the Management of Uterine Fibroids in Canada: Protocol for a Multicenter, Prospective, Noninterventional Study. JMIR Res Protoc. 2018;7(11):e10926. doi:10.2196/10926

4. Senturk M, Polat M, Doğan O, et al. Outcome of Cesarean Myomectomy: Is it a Safe Procedure? Geburtshilfe Frauenheilkd. 2017;77(11):1200–1206. doi:10.1055/s-0043-120918

5. Huang Y, Ming X, Li Z. Feasibility and safety of performing cesarean myomectomy: a systematic review and meta-analysis. J Matern Neonatal Med. 2022;35(13):2619–2627. doi:10.1080/14767058.2020.1791816

6. Goyal M, Dawood AS, Elbohoty SB, et al. Cesarean myomectomy in the last ten years; A true shift from contraindication to indication: A systematic review and meta-analysis. Eur J Obstet Gynecol Reprod Biol. 2021;256:145–157. doi:10.1016/j.ejogrb.2020.11.008

7. Incebiyik A, Hilali NG, Camuzcuoglu A, Vural M, Camuzcuoglu H. Myomectomy during cesarean: a retrospective evaluation of 16 cases. Arch Gynecol Obstet. 2014;289(3):569–573. doi:10.1007/s00404-013-3019-1

8. Sparić R, Malvasi A, Kadija S, Babović I, Nejković L, Tinelli A. Cesarean myomectomy trends and controversies: an appraisal. J Matern Neonatal Med. 2017;30(9):1114–1123. doi:10.1080/14767058.2016.1205024

9. Dedes I, Schäffer L, Zimmermann R, Burkhardt T, Haslinger C. Outcome and risk factors of cesarean delivery with and without cesarean myomectomy in women with uterine myomatas. Arch Gynecol Obstet. 2017;295(1):27–32. doi:10.1007/s00404-016-4177-8

10. El-refaie W, Hassan M, Abdelhafez MS. Myomectomy during cesarean section: A retrospective cohort study. J Gynecol Obstet Hum Reprod. 2020;49(10):101900. doi:10.1016/j.jogoh.2020.101900

11. Sparić R, Papoutsis D, Bukumirić Z, et al. The incidence of and risk factors for complications when removing a single uterine fibroid during cesarean section: a retrospective study with use of two comparison groups. J Matern Neonatal Med. 2020;33(19):3258–3265. doi:10.1080/14767058.2019.1570124

12. Topçu HO, İskender CT, Timur H, Kaymak O, Memur T, Danışman N. Outcomes after cesarean myomectomy versus cesarean alone among pregnant women with uterine leiomyomas. Int J Gynecol Obstet. 2015;130(3):244–246. doi:10.1016/j.ijgo.2015.03.035

13. Kumar R R. The Utility of Caesarean Myomectomy as a Safe Procedure: A Retrospective Analysis of 21 Cases with Review of Literature. J Clin DIAGNOSTIC Res. Published online 2014. doi:10.7860/JCDR/2014/8630.4795

14. Akkurt MO, Yavuz A, Eris Yalcin S, et al. Can we consider cesarean myomectomy as a safe procedure without long-term outcome? J Matern Neonatal Med. 2017;30(15):1855–1860. doi:10.1080/14767058.2016.1228057

15. Özcan A, Kopuz A, Turan V, et al. Cesarean myomectomy for solitary uterine fibroids: Is it a safe procedure? Polish Gynaecol. 2016;87(01):54–58. doi:10.17772/gp/57833

16. Kwon DH, Song JE, Yoon KR, Lee KY. The safety of cesarean myomectomy in women with large myomas. Obstet Gynecol Sci. 2014;57(5):367. doi:10.5468/ogs.2014.57.5.367

17. Ahmadzia HK, Phillips JM, James AH, Rice MM, Amdur RL. Predicting peripartum blood transfusion in women undergoing cesarean delivery: A risk prediction model. PLoS One. 2018;13(12):e0208417. doi:10.1371/journal.pone.0208417

18. Sparić R, Kadija S, Stefanović A, et al. Cesarean myomectomy in modern obstetrics: More light and fewer shadows. J Obstet Gynaecol Res. 2017;43(5):798–804. doi:10.1111/jog.13294

19. Akbas M, Mihmanli V, Bulut B, Temel Yuksel I, Karahisar G, Demirayak G. Myomectomy for intramural fibroids during caesarean section: A therapeutic dilemma. J Obstet Gynaecol (Lahore). 2017;37(2):141–145. doi:10.1080/01443615.2016.1229272

20. Lee Y-E, Park S, Lee K-Y, Song J-E. Risk factors based on myoma characteristics for predicting postoperative complications following cesarean myomectomy. PLoS One. 2023;18(3):e0280953. doi:10.1371/journal.pone.0280953

21. Sei K, Masui K, Sasa H, Furuya K. Size of uterine leiomyoma is a predictor for massive haemorrhage during caesarean delivery. Eur J Obstet Gynecol Reprod Biol. 2018;223:60–63. doi:10.1016/j.ejogrb.2018.02.014

22. Miazga E, Benlolo S, Robertson D, Nensi A. The Use of Uterine Tourniquet to Decrease Blood Loss and Transfusion Rate during Myomectomy: A Systematic Review and Meta-Analysis. J Minim Invasive Gynecol. 2021;28(11):S148–S149. doi:10.1016/j.jmig.2021.09.293

23. Hickman LC, Kotlyar A, Shue S, Falcone T. Hemostatic Techniques for Myomectomy: An Evidence-Based Approach. J Minim Invasive Gynecol. 2016;23(4):497–504. doi:10.1016/j.jmig.2016.01.026

24. Kongnyuy EJ, Wiysonge CS. Interventions to reduce haemorrhage during myomectomy for fibroids. Cochrane Database Syst Rev. 2014;2015(7). doi:10.1002/14651858.CD005355.pub5

